# Genomic analysis of SARS-CoV-2 variants of concern identified from the ChAdOx1 nCoV-19 immunized patients from Southwest part of Bangladesh

**DOI:** 10.1101/2021.08.09.21261778

**Authors:** Hassan M. Al-Emran, Md. Shazid Hasan, Md. Ali Ahsan Setu, Md. Shaminur Rahman, ASM Rubayet Ul Alam, Shovon Lal Sarkar, Md. Tanvir Islam, Mir Raihanul Islam, Mohammad Mahfuzur Rahman, Ovinu Kibria Islam, Iqbal Kabir Jahid, M. Anwar Hossain

## Abstract

**Introduction:** Bangladesh introduced ChAdOx1 nCoV-19 since February, 2021 and in six months, only a small population (3.5%) received their first dose of vaccination like other low-income countries. The remaining populations are struggling with increased rate of infection due to beta and delta variants. Although this uncontrolled COVID-19 pandemic did not leave even the immunized group because of immune escaping capacity of those new variants.

**Methods:** A total of 4718 nasopharygeal samples were collected from 1^st^ March until 15^th^ April, 2021, of which, 834 (18%) were SARS-CoV-2 positive. Randomly generated 135 positive cases were selected for telephone interview and 108 were available and provided consent. The prevalence of SARS-CoV-2 variants and disease severity among both immunized and unimmunized group was measured. A total of 63 spike protein sequence and 14 whole genome sequences were performed from both groups and phylogenetic reconstruction and mutation analysis were compared.

**Results:** A total of 40 respondents (37%, N=108) received single-dose and 2 (2%) received both doses of ChAdOx1 nCoV-19 vaccine which significantly reduce dry cough, loss of appetite and difficulties in breathing compared to none. There was no significant difference in hospitalization, duration of hospitalization or reduction of other symptoms like running nose, muscle pain, shortness of breathing or generalized weakness between immunized and unimmunized group. Spike protein sequence assumed 21 (87.5%) B.1.351, one B.1.526 and two 20B variants in immunized group compared to 27 (69%) B.1.351, 5 (13%) B.1.1.7, 4 (10%) 20B, 2 B.1.526 and one B.1.427 variant in unimmunized group. Those variants were further confirmed by 14 whole genome sequence analysis. Complete genome analysis included seven B.1.351 Beta V2, three B.1.1.7 Alpha V1, one B.1.526 Eta and rest three 20B variant.

**Conclusion:** Single dose of ChAdOx1 couldn’t prevent the new infection or disease severity by the COVID-19 variants of concern, B.1.351, in Bangladesh.

## INTRODUCTION

COVID-19 pandemic continued at a cost of 4.2 million deaths from December 2019 until July 2021. Oxford-AstraZeneca (ChAdOx1 nCoV-19), Pfizer-BioNTech (BNT162b2), Johnson & Johnson (Ad26.COV2.S), Sputnik V (Gam-COVID-Vac) and few other COVID-19 vaccines were administered in different countries. Those vaccines are claiming 67%-100% efficacy in preventing COVID-19 clinical cases [1,2]. Although continuous emergence of new variants introduced debates in preventing the disease. In Bangladesh, the Directorate General of Health Services, announced a total of 3,227,598 samples were tested, of which, 513,510 were positive and 7,559 (1.47%) died (https://corona.gov.bd/storage/press-releases/December2020/KsvQiLw3eIdbv2GTq8RR.pdf) until December 2020. In 2021 until August, the number of tests increased into more than double (N=7,790,423) with 1,264,328 SARS-CoV-2 positive cases and 20,916 deaths (1.67%) (http://dashboard.dghs.gov.bd/webportal/pages/covid19.php). The increased death-case ratio attributed due to B.1351 [3] and B1.617.2 [4] variants . However, the number was substantially underestimated due to limited transportation towards the diagnostic facilities in many regional parts of Bangladesh. Before the second wave of the pandemic, Bangladesh has introduced ChAdOx1 nCoV-19 and until 4th of August, only 2.7% received two doses and 3.1% received one dose of vaccination, (https://ourworldindata.org/covid-vaccinations?country=OWID_WRL). Therefore, a large proportion of the population remain unimmunized or received only the first dose of ChAdOx1 nCoV-19 vaccine in Bangladesh. Almost 2.5 billion doses of different vaccines have been distributed among 21% of the global population although only 0.8% population in low-income countries received only one dose of a vaccine (https://ourworldindata.org/covid-vaccinations?country=OWID_WRL). Several studies have been performed to evaluate the safety and efficacy of first doses vaccines [5]. First dose of the ChAdOx1 nCoV-19 was found to be effective against 63.9% of symptomatic and asymptomatic cases [5]. In another randomised controlled trials, one standard dose participants showed 64.1% vaccine efficacy [1]. Bernal et al.(2021) reported that first dose ChAdOx1-S effectiveness reached 60% from 28-34 days with further increases to 73% from 35 days to onward for B.1.1.7 variant [6].

A large number of people are being reported to be infected with SARS-CoV-2 during this interim period with those immune-escape variants. Continuous genetic surveillance would assist to reveal those new variants capable of escaping the current immunization. This study emphasized on the infected cases after immunization and analyze the variants of SARS-CoV-2.

## METHOD

### Ethical Approval

This study is approved by the ERC of Jashore University of Science and Technology (ERC no: ERC/FBST/JUST/2020-51). Verbal consent was taken from all participants after reading the purpose and objectives. Positive respondents were then asked for a telephone interview with the study questionnaire and enroll the respondent in this study.

### Study population

This study was conducted from 1^st^ March until 15^th^ April, 2021 in South-West part of Bangladesh. The Genome Centre, Jashore University of Science and Technology, Jashore, Bangladesh is conducting COVID-19 diagnosis and covering four districts (Jashore, Magura, Narail and Jhenaidha) as a part of the national surveillance system approved by DGHS [7].

A total of 4718 nasopharygeal samples were collected from 3 districts (Jashore, Narail and Magura) and tested for SARS CoV-2. The RNA was released from the samples using QuickExtract™ RNA extraction kit (Lucigen, Wisconsin, USA) following the manufacturer’s instruction. Then, 10 µL of each viral RNA extract was amplified by one-step real-time reverse-transcriptase polymerase chain reaction (RT-PCR) using Novel Coronavirus Nucleic Acid Diagnostic Kit (Sansure Biotech Inc., China). A total of 834 (18%, N=4718) samples were identified for the presence of N-gene or orf1b gene with a CT-value <40 and interpreted as SARS-CoV-2 positive according to the manufacturer’s instruction. The participants were excluded from the study if they were unable to participate or receive our calls or does not have the vaccine card or have the history of immunological disorder. In this study, we included day-wise random number generated 135 cases (out of 834), of which, only 108 were available for telephone interview with a prior verbal consent. All respondents (n=108) were interviewed with a pre-tested structured questionnaire over the phone. In the case of unavailability of the respondents due to death, first kin attended the interview.

### Vaccination status

All the participants were asked about their vaccination status, number of doses and date of vaccination. The vaccination status was confirmed by vaccine card provided by DGHS, Bangladesh (https://surokkha.gov.bd/vaccine-status) or the message provided by the DGHS.

### Statistical analysis

Descriptive analysis was conducted using frequency distribution to explore the sociodemographic profile, COVID-19 related experience and information about hospitalization. Chi-square test of independence was performed to explore the relationship between vaccination status and respondents’ characteristics. In terms of smaller sample size, fisher exact tests were used to determine the association between the outcome and disaggregated variable.

### Spike protein sequencing

RNA was extracted from 140 µl of left-over samples using the QIAamp Viral RNA Mini Kits (QIAGEN, USA) according to the manufacturer protocol. PCR was performed with 7µl of RNA extract using the Luna® Universal One-Step RT-qPCR Kit (New England Biolabs Inc., USA) with the primers for Rbd region of the spike portion of the genome. The details of the protocol were shown in the supplementary Table S1 [8]. The amplicons, confirmed in 1% (w/v) agarose, were sequenced using the BigDye® Terminator v3.1 Cycle Sequencing Kit (Applied Biosystem, ThemoFisher Scientific, Inc., USA). The sequenced FASTA files were initially cleaned with Chromas Pro (https://technelysium.com.au/wp/) and aligned with the reference sequence (NC_045512.2/SARS-CoV-2/Wuhan-Hu-1) using Molecular Evolutionary Genetics Analysis (MEGA X) software [9] to detect the presence of any mutations.

### Spatial distribution mapping

Spatial distribution of COVID-19 cases was mapped using the ArcGIS (version 10.6) software [10]. The nearest geolocations of the COVID-19 cases were approximated with the help of Google Earth Pro platform (a freely available google product, formerly known as Keyhole Earth Viewer) and as per the description of the respondents.

### Whole genome sequencing

First-strand cDNAs were prepared from the extracted viral RNA using SuperScript™ III First-Strand Synthesis System (Invitrogen™,Thermo Fisher Scientific, USA). The concentrations of cDNA were determined using the dsDNA HS Assay Kit with Qubit 4 Fluorometer (Thermo Fisher Scientific, USA). The Ion AmpliSeq™ SARS-CoV-2 Research Panel (Thermo Fisher Scientific, USA) was used to prepare Ion AmpliSeq™ libraries. Two 5X primer pools that target 237 amplicons specific to the SARS-CoV-2 was used during target amplification PCR and number of amplification cycles was determined based on their viral copy number. Each amplified samples were subjected to 2µL FuPa reagents for partial digestion. Digested amplicons were ligated with P1 adapter and Barcode Adapters (Ion Torrent™, Thermo Fisher Scientific, USA). Magnetic bead cleanup was performed using Agencourt™ AMPure™ XP Reagent (Beckman Coulter, USA) to purify barcoded Ion AmpliSeq™ libraries. The library concentration was calculated using Ion Library TaqMan® Quantitation Kit and diluted into 100 picomolar (pM) according to the manufacturer’s protocol. All equimolar libraries were pooled for the preparation of template-positive Ion Sphere™ Particles (ISPs) using the Ion 530™ Kit – OT2 (Thermo Fisher Scientific, USA) on the Ion One Touch™ 2 System. Template-positive ISPs were enriched on Ion One Touch™ ES system and loaded in Ion 530™ chip with control ISPs for sequencing into Ion S5™ System.

### SARS-CoV-2 Genome consensus generation

Short read of Binary Alignment Map files were converted to Fastq file by using SAMtools v1.12 [11] and the quality checked by using FastQC 0.11.9 [12]. Trimmomatic v0.39 [13] was used to trimmed low quality read (phred score less than 20) and consensus were generated by using Burrows-Wheeler Aligner (BWA v0.7.17) [14], SAMtools v1.12 [11] and BEDTools v2.30.0 [15] by aligning a reference genome (hCoV-19/Wuhan/WIV04 (GenBank accession no. MN996528.1). Further indel and area wise mutation coverage were checked by using Snippy [16] and corrected the read.

### Phylogenetic reconstruction and mutation analysis

Nextstrain SARS-CoV-2 phylogetetic tree generation pipeline (https://nextstrain.org/sars-cov-2) has been used to reconstruct the phylogenetic tree with Nextstrain clade of the sequenced (n=14) [17]. Nextstrain uses MAFFT [18] as an alignment tools, IQ-TREE [19] and TreeTime [20] to reconstruct tree and Auspice web server (https://auspice.us/) to visualize the tree. Mutations and clades have assigned from CoVserver mutation app (https://www.gisaid.org/epiflu-applications/covsurver-mutations-app/) of Global initiative on sharing all influenza data (GISAID) where hCoV-19/Wuhan/WIV04 (GenBank accession no. MN996528.1) was used as a reference genome. Heatmap with nonsynonymous mutations was generated by using an R package Pheatmap [21] and violin plot of between synonymous and nonsynonymous mutations was generated by using ggplot2 [22]. The Pearson correlation coefficient was measured between the immunized and unimmunized group.

## RESULTS

### Description of study population

This study reports information of 108 COVID-19 positive cases tested between March 1 to April 15, 2021. The socio-demographic profiles, co-morbidity, vaccination and COVID-19 associated symptoms, diagnosis and treatment information of the studied population are presented in Table 1. Studied respondents were dominated by males (75%) and were mainly from aged group 50-60 years (34%) followed by 40-50 years (28%). The mean age of the respondents were 50 years (maximum 76.4, minimum 14.4 and median 50.5 years). Estimated Body Mass Index (BMI) suggests that half of the studied respondents can be characterized as either overweight (40.7%) or obese (10.2%). Majority of the respondents (66.4%) completed their secondary or higher education, whereas 14% of them didn’t complete their primary education. Occupationally, they were engaged in service or business (60.2%) homemaker (18.5%) and unemployed or retired (13.0%).

**Table 1:** Statistical analysis of the study patients’ profile.

| Variables | Section I: All ages groups |  |  | Section II: 16 to 70 years |  |  |
| --- | --- | --- | --- | --- | --- | --- |
|  | 1st dose | Non immunized | P value | 1st dose | Non immunized | P value |
|  | % (n) | % (n) |  | % (n) | % (n) |  |
| Sociodemographic profile |  |  |  |  |  |  |
| Sex |  |  |  |  |  |  |
| Male | 83.3 (35) | 69.7 (46) | 0.111 | 82.5 (33) | 70.5 (43) | 0.171 |
| Female | 16.7 (7) | 30.3 (20) |  | 17.5 (7) | 29.5 (18) |  |
| BMI |  |  |  |  |  |  |
| Normal | 45.2 (19) | 51.5 (34) | 0.777 | 42.5 (17) | 50.8 (31) | 0.731 |
| Overweight | 45.2 (19) | 37.9 (25) |  | 47.5 (19) | 39.3 (24) |  |
| Obese | 9.5 (4) | 9.5 (7) |  | 10.0 (4) | 9.9 (6) |  |
| Education |  |  |  |  |  |  |
| No education or primary incomplete | 4.8 (2) | 20.0 (13) | 0.009 | 5.0 (2) | 18.0 (11) | 0.027 |
| Secondary incomplete | 11.9 (5) | 24.6 (16) |  | 12.5 (5) | 24.6 (15) |  |
| Secondary or higher | 83.3 (35) | 55.4 (36) |  | 82.5 (33) | 57.4 (35) |  |
| Occupation |  |  |  |  |  |  |
| Unemployed | 14.3 (6) | 12.1 (8) | 0.223 | 12.5 (5) | 9.8 (6) | 0.440 |
| Service/business | 69.1 (29) | 54.6 (36) |  | 70.0 (28) | 59.0 (36) |  |
| Homemaker | 14.3 (6) | 21.2 (14) |  | 15.0 (6) | 21.3 (13) |  |
| Others | 2.4 (1) | 12.1 (8) |  | 2.5 (1) | 9.8 (6) |  |
| Presence of any comorbidities |  |  |  |  |  |  |
| Yes | 59.5 (25) | 43.9 (29) | 0.114 | 60.0 (24) | 44.3 (27) | 0.122 |
| No | 40.5 (17) | 56.1 (37) |  | 40.0 (16) | 55.7 (34) |  |
| Comorbidities (Multiple response) |  |  |  |  |  |  |
| Diabetes | 19.1 (8) | 30.3 (20) | 0.193 | 17.5 (7) | 29.5 (18) | 0.171 |
| High blood pressure | 31.9 (13) | 31.0 (21) | 0.925 | 30.0 (12) | 32.8 (20) | 0.768 |
| Clinical manifestation |  |  |  |  |  |  |
| Symptoms (Multiple response) |  |  |  |  |  |  |
| Fever | 50.0 (21) | 67.7 (44) | 0.067 | 47.5 (19) | 66.7 (40) | 0.058 |
| Dry cough | 31.0 (13) | 51.5 (34) | 0.036 | 30.0 (12) | 49.2 (30) | 0.056 |
| Running nose | 7.1 (3) | 13.6 (9) | 0.361 | 7.5 (3) | 14.8 (9) | 0.355 |
| Muscle pain | 26.2 (11) | 19.7 (13) | 0.429 | 25.0 (10) | 19.7 (12) | 0.526 |
| Difficulties in breathing | 7.1 (3) | 27.3 (18) | 0.012 | 7.5 (3) | 27.9 (17) | 0.012 |
| Loss of smell | 19.1 (8) | 30.3 (20) | 0.193 | 17.5 (7) | 31.2 (19) | 0.125 |
| Loss of taste | 21.4 (9) | 25.8 (17) | 0.608 | 22.5 (9) | 27.9 (17) | 0.546 |
| Loss of appetite | 7.1 (3) | 22.7 (15) | 0.034 | 5.0 (2) | 23.0 (14) | 0.016 |
| Complications after covid 19 positive (Multiple response) |  |  |  |  |  |  |
| Generalized weakness | 11.9 (5) | 18.2 (12) | 0.383 | 12.5 (5) | 19.7 (12) | 0.346 |
| Emotional changes | 7.1 (3) | 7.6 (5) | 1.000 | 7.5 (3) | 8.2 (5) | 1.000 |
| Dementia | 9.5 (4) | 6.1 (4) | 0.709 | 10.0 (4) | 6.6 (4) | 0.709 |
| COVID-19 severity |  |  |  |  |  |  |
| Hospitalized | 11.9 (5) | 18.2 (12) | 0.383 | 10.0 (4) | 14.8 (9) | 0.485 |
| Duration |  |  |  |  |  |  |
| No hospitalization | 88.1 (37) | 81.5 (53) |  | 90 (36) | 85 (51) |  |
| 1 week | 7.1 (3) | 10.8 (7) | 0.780 | 7.5 (3) | 10 (6) | 0.723 |
| More than 1 week | 4.8 (2) | 7.7 (5) |  | 2.5 (1) | 5 (3) |  |
| Oxygen | 9.5 (4) | 15.2 (10) | 0.396 | 7.5 (3) | 14.8 (9) | 0.355 |
| **Spike protein variants** |  |  |  |  |  |  |
| Sanger variant |  |  |  |  |  |  |
| B.1.351 | 52.4 (22) | 39.4 (26) | 0.185 | 52.5 (21) | 39.3 (24) | 0.193 |
| B.1.1.7 | 0.0 (0) | 7.6 (5) | 0.154 | 0.0 (0) | 4.9 (3) | 0.275 |
| B.1.526, B.1.427 and Conventional | 7.1 (3) | 10.6 (7) | 0.737 | 7.5 (3) | 9.8 (6) | 1.000 |
| Unknown | 42.4 (17) | 40.5 (28) | 0.841 | 40.0 (16) | 45.9 (28) | 0.559 |

The study reported several co-morbidities among half of the respondents. Most often the respondents reported high blood pressure (31.5%) followed by diabetes (26%), cardiac diseases (9.3%) and asthma (4.6%) etc. Majority of the respondents suffered from fever (61%), dry cough (43.5%), loss of smell (26%), loss of taste (24%) and muscle pain (22%) during the infection. The respondents approached for a COVID-19 test due to their generalized sickness (79.6%), exposure with COVID-19 patient (8.3%) and other reasons (12%). About 63% respondents reported a complete recovery, while 30.6% were still suffering (generalized weakness, emotional changes, and dementia) and 6.5% died (Table S2). About 39% (n = 42) of the respondents were infected after the COVID-19 vaccination. First dose of ChAdOx1 nCoV-19 vaccine was received by 40 (37.0%) cases and both doses were completed by only 2 (2%) cases. The average duration between vaccination (partially or completely immunized) and COVID 19 diagnosis was 32(±17) days. Almost 86% and 69% of the respondents were infected minimum after 2 and 3 weeks of administration of first dose, respectively (Figure 1). Among the respondents, 15.7% (n = 17) suffered from severe consequences and required hospitalization for 1 to 17 days (mean 7.72 days and median 6 days). Additional oxygen was supplemented to 13% (N=14) of the respondents during their hospitalization, of them, 7 respondents (6.5%) were shifted to Intensive care unit (ICU) and 4 died. Three respondents died at home before rushing towards the hospital.

**Figure 1:**
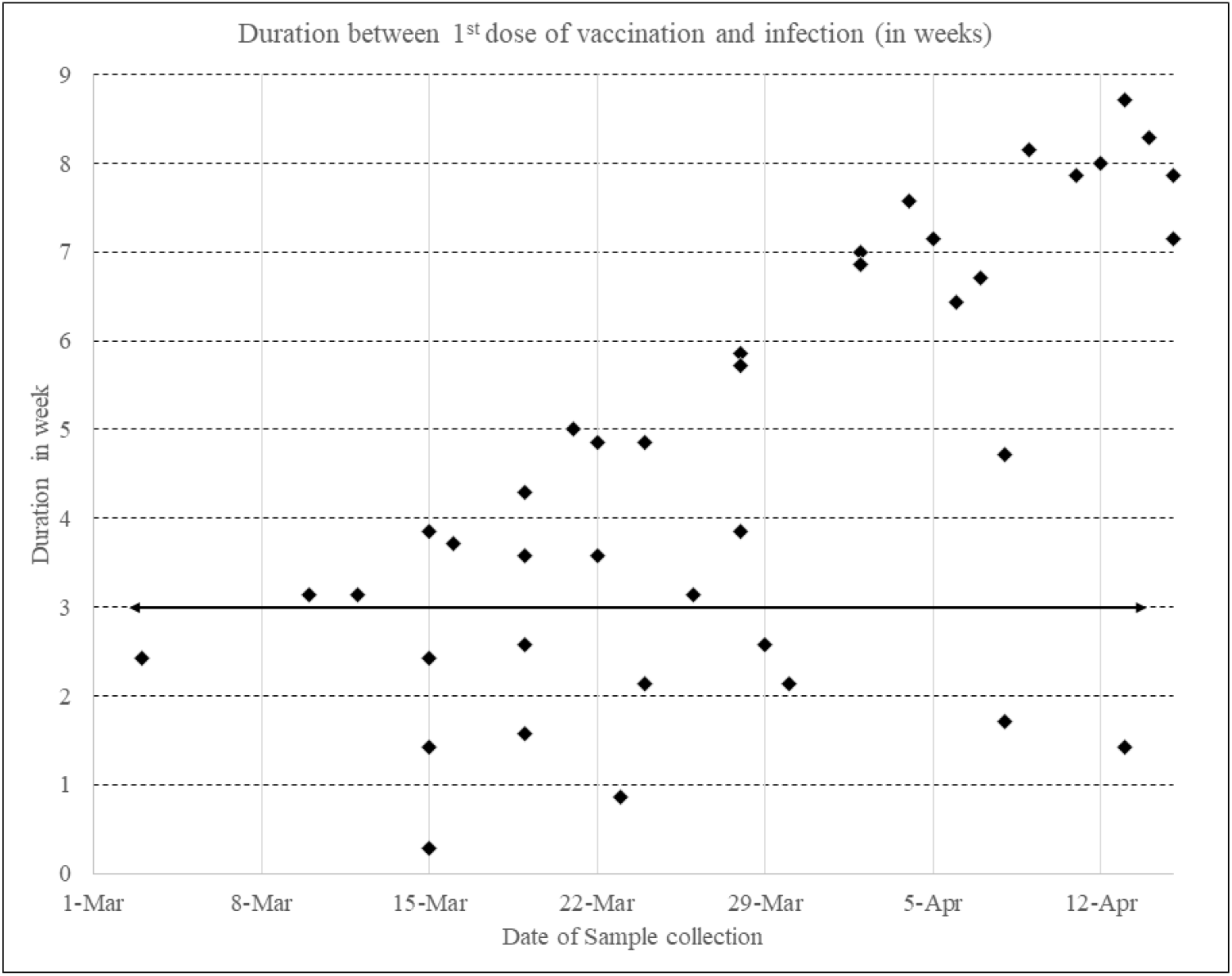
Duration between SARS-CoV-2 infection and first dose of immunization. 86% (N=36) of the cases were infected after 14 days and 71% (N=30) of them were infected after 21 days of vaccination. Red dots represent the death cases and black dots represent the survived cases.

We performed the comparative analysis of vaccination status and respondents’ characteristics (Table 1). We have disaggregated the analysis in two sections, section I considering all age groups, section II considering only 16 to 70 years old respondents. In all age group, we observe that education status has significant relationship with the vaccination status. Most of respondents (83.3%) immunized group completed their secondary or higher education. Difficulty of breathing and loss of appetite were significantly higher in the unimmunized group (27% vs. 7%, and 23% vs. 7%, respectively, p<0.05) which was also found among 16-70 years. Dry cough was also significantly high in all ages among unimmunized group (p<0.05). Fever was more common among in that group although not significant (p=0.067). Apparently, in our study we couldn’t found any significant difference of hospitalization (p=0.383) (Figure S1) and duration of hospitalization (p=0.780). Hospitalization rate of comorbid patients were also similar (24% both) between both groups (Table S3). Additionally, we compared the mean CT values of immunized and unimmunized patients, where we identified a significant increase of *N*-genes ((p=0.026) and Orf-1b gene (p =0.052) among unimmunized (Figure S2).

### Spatial distribution of identified COVID-19 variants

The spatial distribution of COVID-19 variants suggested a clear clustering and localized proliferation (Figure 2). South African variants (B.1.351) showed prominent clusters and, thus results localized hotspots in the Jashore Municipality.

**Figure 2:**
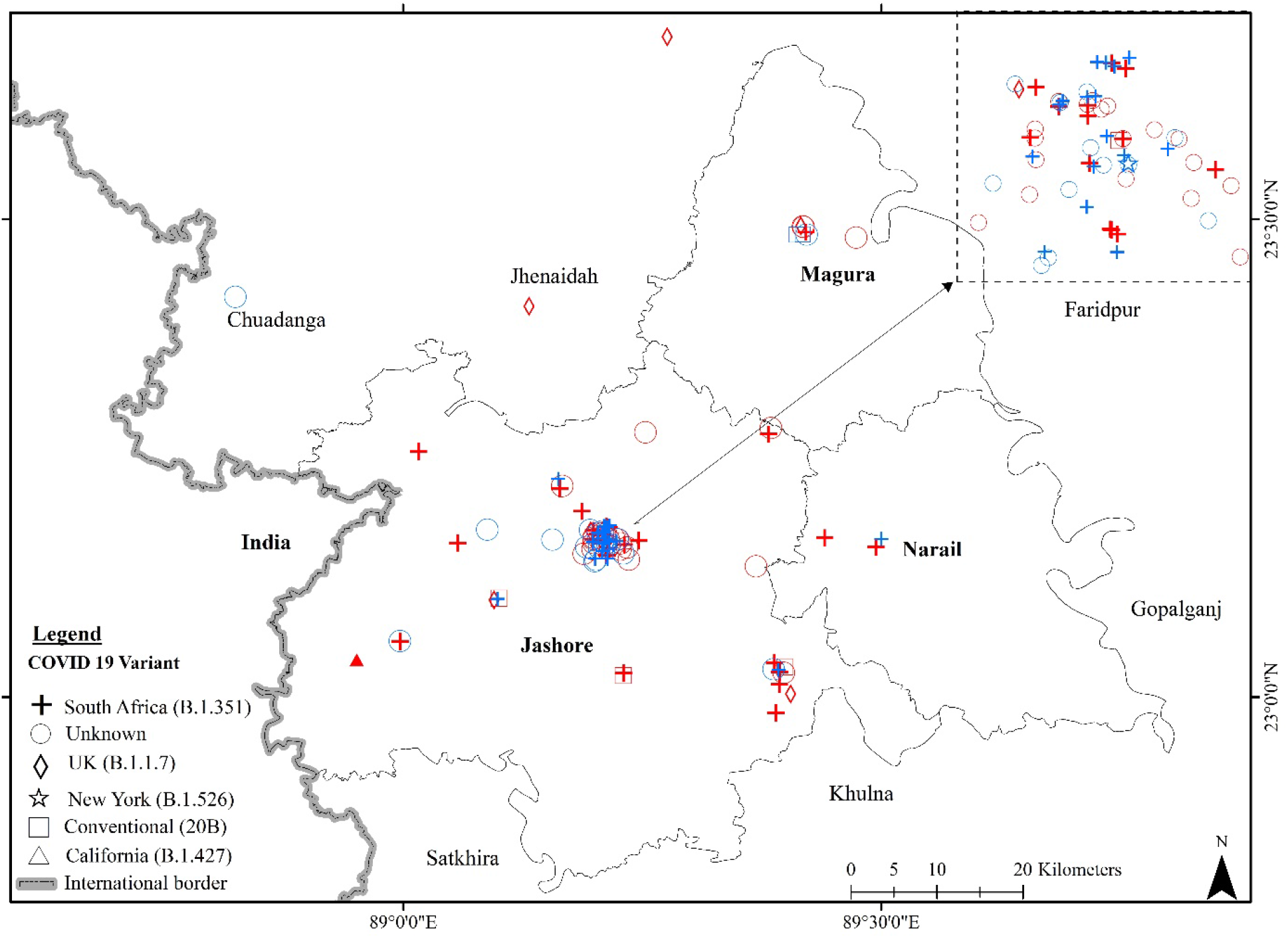
Geospatial location of SARS-CoV-2 variants. Red marks indicate the unimmunized SARS-CoV-2 infected patients. Blue marks indicate the partial or complete immunized COVID-19 patients.

### Spike protein variants

In this study, only 78 samples had the threshold cycle (CT value) below 35 in RT-PCR screening test and considered as potential candidate for partial or complete genome sequencing, of which, only 76 left-over samples were available. The concentration of spike protein RNA might be low in rest 13 samples and could not be visualized in gel electrophoresis. The template RNA provided 850bp spike protein amplicons in 63 patients’ samples, of which, 24 patients were partially or completely immunized before the COVID-19 infection. Spike protein sequence mutation analysis revealed that 21 (87.5%) strains might be B.1.351 variant, one was B.1.526 variant and two were conventional without any mutation. Among the unimmunized 39 viral strains, spike protein sequences analysis suspected 27 (69%) B.1.351, 5 (13%) B.1.1.7, 4 (10%) conventional, 2 B.1.526 and one B.1.427 variant. Figure 2 showed the spatial distribution of 63 SARS-CoV-2 variants among both groups, rest 45 out of 108 were marked as unknown in the map. Most cases (N=95) were detected from the largest district, Jashore. Few other cases were located in Magura (N=10) and Narail (N=3). Mutation analysis revealed that 16% (10 out of 63) of the strains had non-synonymous unique mutations in their RBD region of the spike protein; 7 (18%, n=39) of which belongs to unimmunized groups and 3 (12.5%, n=24) belong to immunized group. Three B.1.351 variants and 2 B.1.526 variants each contain single unique mutation and two others B.1.351 variants contain multiple unique mutation among unimmunized group (Table 2). Compared to that, immunized group contain F515S mutation in two and W353R in one B.1.351 variant.

**Table 2:** Identification of SARS-CoV-2 variants and their non-synonymous unique mutations.

| **Spike protein sequences** |  |  |  |  |
| --- | --- | --- | --- | --- |
| Variant | Immunized<br>(number of variants) | Unique Mutations<br>(No of strains) | Unimmunized<br>(number of variants) | Unique Mutations (No of strains) |
| B.1.351 | 21 | F515S (2)<br>W353R (1) | 27 | -I402N (1)<br>-I402M (1)<br>-G413W (1)<br>-N394K, E406K, D405E, Q414E, S375F (1)<br>-N394K, E406K, Q414E, S375F (1) |
| B.1.1.7 | 0 |  | 5 |  |
| Conventional | 2 |  | 4 |  |
| B.1.427 | 0 |  | 1 |  |
| B.1.526 | 1 |  | 2 | -F377I (1), -Q414E (1) |
| **Whole genome sequences** |  |  |  |  |
| Variant | Immunized<br>(frequency of unique mutation in a single strain) | Average Unique Mutations | Unimmunized<br>(frequency of unique mutation in a single strain) | Average Unique Mutations |
| B.1.351 | 11, 22, 19, 21 | 18.3 | 20,22,11 | 17.7 |
| B.1.526 | 13 |  | 22 |  |
| B.1.1.7 |  |  | 34 |  |
| Conventional/<br>Unrecognized (20B) | 7, 13 | 10 | 22, 12 | 17 |

### Complete genome and mutation analysis

The complete genome of SARS-CoV-2 analyzed to map a radial phylogenetic tree (Figure 3) and it has found that six different variants for both immunized and unimmunized groups (Table 2 and Table S4). Complete genome analysis (n = 14) included 7 B.1.351 Beta V2 variant, 3 B.1.1.7 Alpha V1 variant, one B.1.526 Eta variant and rest three 20B variant (Figure 4) from both groups. The t-test found a non-significant difference between the mean frequency of synonymous (p=0.396) and non-synonymous (p=0.083) mutations between the immunized and unimmunized group (Figure 5). N501Y mutation was found in 9 strains; 3 harbored in partially immunized patients and 6 harbored in unimmunized patients. P681H mutation were possessed by 3 strains and all were belonged to unimmunized patients. H69–V70 deletion found in 3 strains (2 unimmunized, 1 partially immunized patients) and Y144 deletion found in 2 strains (1 unimmunized, 1 partially immunized patients). E484K mutation was possessed by 7 (50%) strains among which 4 patients were partially immunized and 3 patients were unimmunized. K417N mutation was found in 4 partially immunized patients and 2 unimmunized patients. L452R substitution is found in 2 strains (Figure 4).

**Figure 3:**
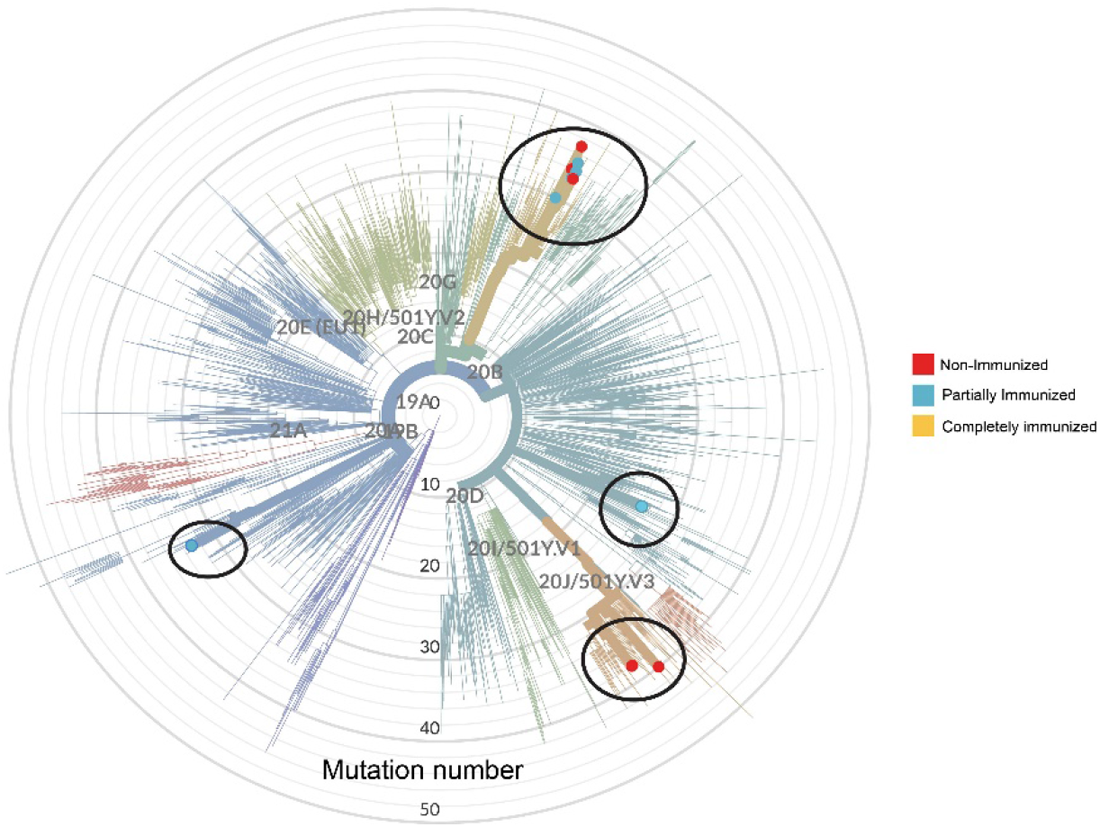
Phylogenetic of tree with their existing clade with world reference genome. Red color represents unimmunized variants, sky-blue represents partially immunized and orange color represents completely immunized variants in the tree.

**Figure 4:**
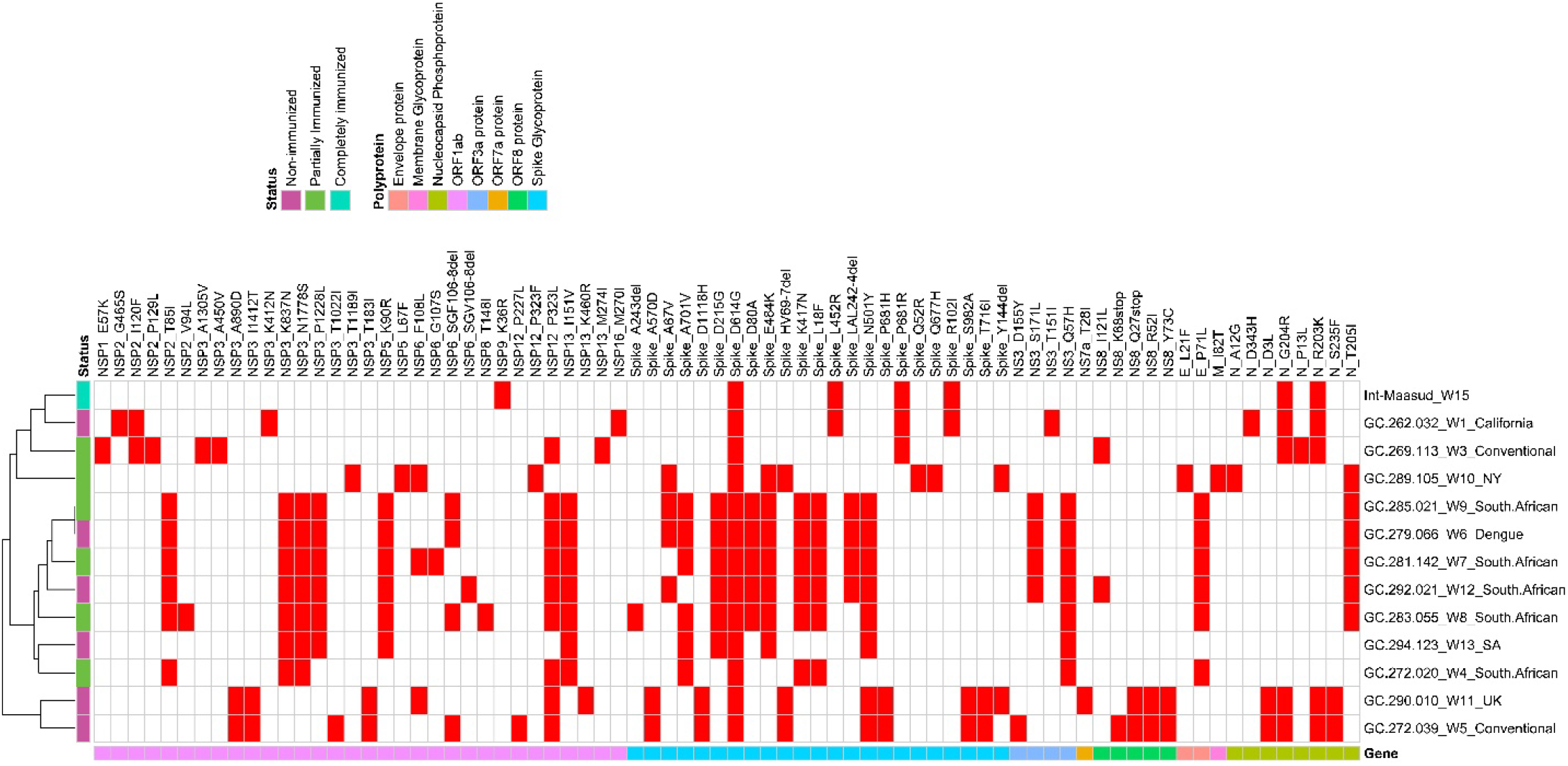
Comparison of amino acid substitutions retrieved from genome sequences of SARS-CoV-2 viruses among immunized and unimmunized patients. Mutations and amino acid substitutions were retrieved from Nextclade (https://clades.nextstrain.org/).

**Figure 5:**
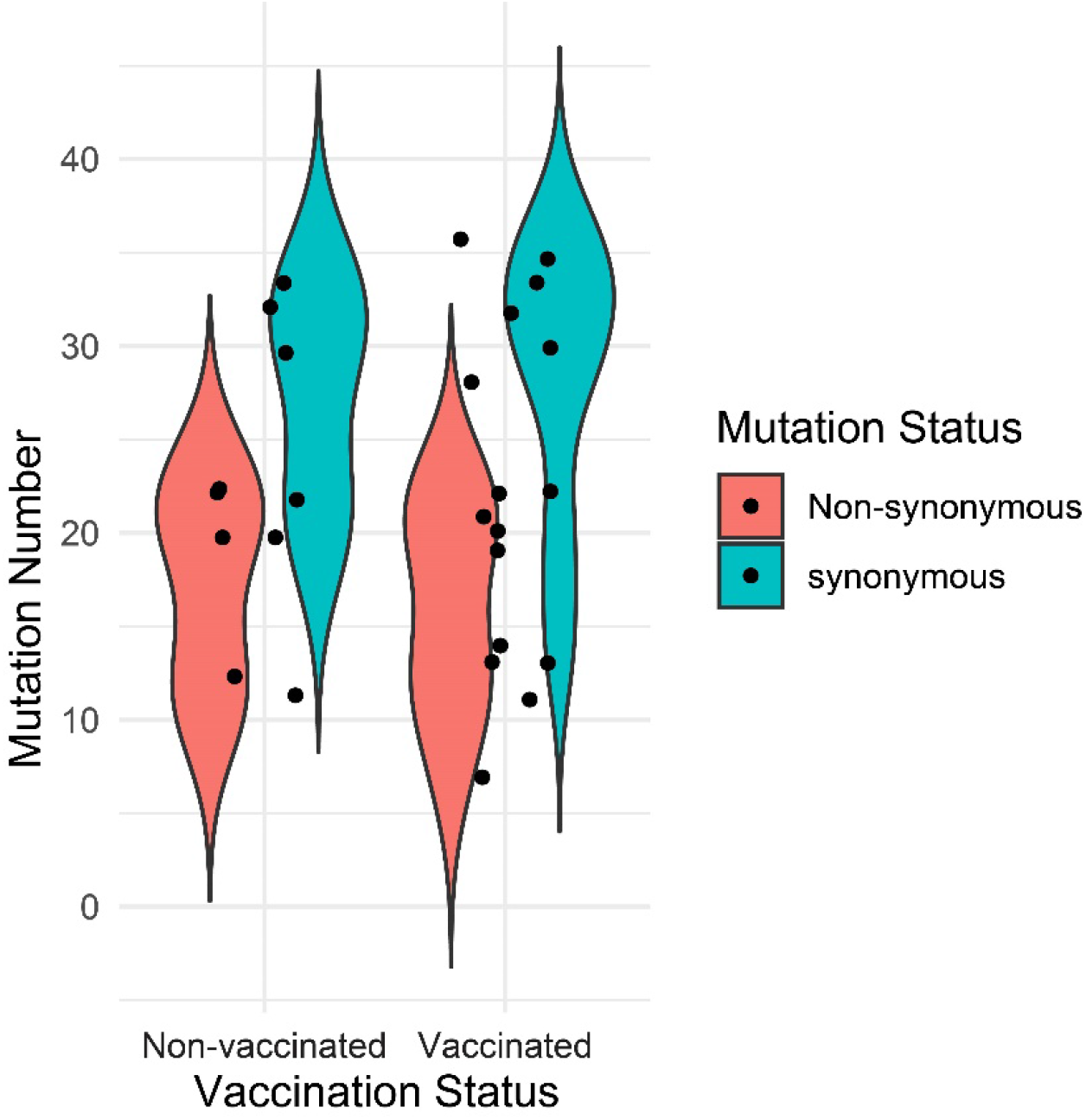
Synonymous (p=0.396) and non-synonymous (p=0.083) mutation status between immunized and unimmunized patients.

## DISCUSSION

Emergence of SARS-CoV-2 variants is a global problem during COVID-19 pandemic [23–26]. Those variants has been designated as variants of interest (VOI) and variants of concern (VOC) by WHO (https://www.who.int/en/activities/tracking-SARS-CoV-2-variants/) and CDC (https://www.cdc.gov/coronavirus/2019-ncov/variants/variant-info.html) depending on its genetic changes that affects transmissibility, disease severity, immune escape, diagnostic or therapeutic escape, increasing prevalence, increasing virulence, decreasing the effectiveness of vaccines etc.

This study found that first or second dose of ChAdOx1 did not prevent the new infections by the variants of interests and variants of concern (B. 1.526, B.1.351, and 20B) in Bangladesh except the B.1.1.7 variant. Most of the viruses (87.5%) detected from the vaccinated patients were identified as B.1.351 variant indicating that the ChAdOx1 couldn’t prevent new infection by this variant of concern (Table 2). The SARS-CoV-2 variants and the efficacy of the different vaccines has been reviewed by Bian et al. (2021) [27] and demonstrated that efficacy was reduced from ranges 0.78 to 9. The highest reduction was 9.0 observed for B.1.351 variant of ChAdOx1 [28]. Madhi et al. also noted that ChAdOx1 vaccine efficacy was only 10.4% after two doses in South Africa where B.1.351 was predominant [29]. Few studies found the similar results that the ChAdOx1 vaccine are not effective against recent variant of concerns such as B.1.617.2 [6] and Delta 1 [30]. That raised a dubious situation about the efficacy COVID-19 vaccine among the population [31].

A study in Argentina, Brazil Chile, Colombia, Mexico, Peru, South Africa and the United States found that single dose ChAdOx1 vaccine efficacy was 52 to 64% [32]. Few studies claimed that single dose vaccination protect from infections including B.1.1.7 [6,33–35]. However, in our study, most of the COVID-19 infections (71%, N=30) were identified after 3^rd^ weeks of vaccine administration, with a mean duration of 32(±17) days (Figure 1). Even after 4^th^ week (52%, N=22) of vaccination the antibody couldn’t prevent new infection by SARS-CoV-2 B.1.351 variant. A study by Tauzin et al. reported that BNT162b2 vaccine was 90% effective after 3 weeks by the production of g anti-receptor binding domain and spike antibodies with Fc-mediated effector functions and cellular CD4+ T cell responses [36].

However, Madhi et al. observed that ChAdOx1 vaccine couldn’t offer any protection against low and mild infection of SARS-CoV-2 due to B.1.351 variant [29]. In our study, we found that the ChAdOx1 vaccine significantly reduce dry cough, loss of appetite and difficulties in breathing. However, there was no significant reduction of other symptoms like, running nose, muscle pain, shortness of breathing, generalized weakness, emotional changes etc (Table 1). Madhi et al couldn’t include any hospitalized cases to observe the effect on severe cases. Our study found no significant differences in disease severity in terms of hospitalization (p=0.383) (Figure S1) or the duration of hospitalization (p=0.780) when compared between the immunized and unimmunized group. Another previous study by Bernal et al. [6] reported 37% reduction of hospitalization in case of single dose vaccination by ChAdOx1, although we found only 6.3% reduction (Table 1). SARS-CoV-2 infected patients comorbidity affected by diabetes, asthma or high blood pressure were facing the severe consequences throughout the pandemic. Previous studies found 28-50% of hospitalization among comorbid patients from January to march 2020 [37]. The rate of hospitalization of comorbid patients in our study was 23.5% among the immunized and 24.1% among the nonimmunized groups (Table S3). In another study observed that one dose vaccination with either BNT162b2 or ChAdOx1 nCoV-19 could reduce the hospitalization rate by 51% among the elderly group aged above 80 [38].

Although there was no significant difference in hospitalization and comorbid groups, the RT-PCR results found a significant difference in threshold cycles values (Figure S2) of immunized and unimmunized groups (p=0.026 for N-gene, p=0.052 for ORF1b gene). A previous study also found the similar results [39] without any correlation of the vaccine efficacy or disease severity and outcomes. However, Pritchard et al showed significant difference of Ct values with reduction of symptomatic infections, hospitalization and death [34].

There were no significant differences found in the number of the synonymous and non-synonymous mutations on the whole genome sequences of viruses collected from immunized and unimmunized patients, even though higher numbers of mutations were observed for unimmunized groups (Figure 5). The unimmunized groups showed more unique mutations in spike protein as well as in other positions. Important mutations that can play vital roles in evading immune responses are the characteristic of variant of concerns. In our study we found those mutations in both immunized and unimmunized groups. Asparagine to tyrosine conversion at 501 amino acid position (N501Y) of spike glycoprotein helps the virus latch on more tightly to human cells. But the mutation is not likely to help the virus evade current vaccines [40]. This mutation harbored in 3 partially immunized patients and 6 unimmunized patients. Proline to Histidine substitution (P681H) at spike glycoprotein may help infected cells create new spike proteins more efficiently (Jonathan Corum, 2021) which was found 3 unimmunized patients in our study. The histidine and valine deletion from 69 and 70 positions (H69–V70) and tyrosine deletion from 144/145 position (Y144/145) in spike glycoprotein alter the shape of the spike and may help it evade some antibodies [40]. H69–V70 deletion found among 3 patients (2 unimmunized, 1 partially immunized) and Y144 deletion found among 2 patients (one in each group). Lysine to asparagine alteration at 417 position (K417N) of spike glycoprotein helps the virus bind more tightly to human cells [40]. This mutation was found in 6 study patients (4 in immunized and 2 in unimmunized patients). E484K substitution may help the virus evade antibodies [40] which was found 4 immunized and 3 unimmunized patients. L452R substitution is common in B.1.427, but not yet shown to be more infectious (Figure 4).

Bangladesh had started pilot vaccination program with ChAdOx1 on January 27, 2021 and nationwide vaccination program started on February 7. This study investigated to observe the variants after the selective pressure with first dose of vaccination in the population (Figure 2) and the samples were collected from 1^st^ March to 15^th^ April, 2021. During that time Bangladesh was facing the second wave of SARS-CoV-2 infection in which Variant B.1.351 was found predominant [24]. The study was conducted before the first case of B.1.617.2 variant were reported in Bangladesh by our research group [41]. Whole genome sequencing is an expensive and time-consuming process; thus, it was not suitable for large-scale variant surveillance in countries with limited research funding like Bangladesh. The possibility and necessity of detecting important mutations by partial sequencing of the viral genome was mentioned by our research group [8,42]. Detection of the ‘signature mutations’ in spike proteins by Sanger sequencing can be useful for the initial screening of the variants. This initial data can be verified by performing whole genome sequences considering the virus type, pathology and itinerary of the infected patient. In this study we applied both partial sequencing and whole genome sequencing for variant screening (Table 2 and Figure 3). Although this study is limited to low number of participants, thus it can provide only a little update on SARS-CoV2 variant profile during the vaccination program in Bangladesh.

This is the first report from Bangladesh about the post vaccination variants during the second wave of pandemic by B.1.351 that provided the impact of selective pressure in the community after the first dose of vaccination. The continuous monitoring of variants is essential considering the large number of populations might receive only single dose for a while and there are limited supplies of this most demanding vaccine. The impact of vaccination should be carefully measured because a large number of populations are getting infected during the interim period between two doses. Those, who get infected with COVID-19 after the first dose of vaccination, might be kept in the least priority list, rather offering new personnel who neither receive any dose nor was recently infected. WHO shall also make a suggestion or guideline for the recently infected personnel regarding the minimum time gap between active infection and next dose of vaccine.

## Data Availability

All data can be found in GISAID.

## Acknowledgments

We would like to acknowledge the Genome Center, Jashore University of Science and Technology, to provide the laboratory facilities to perform this study.

## Data availability statement

The whole genome sequence data analysed in this study were deposited in GISAID (https://www.gisaid.org/) with accession no. mentioned in Table S4.

## Authors contributions

HMA: Conceptualization, methodology, validation, formal analysis, investigation, visualization, writing original draft, funding aquisition.

MSH: Methodology, investigation, resources, validation, writing original draft.

MAAS: Investigation, analysis, data curation.

SR: Sortware, Formal analysis, data curation.

ASMRA: writing review and editing, resources

SLS: Investigation, analysis, data curation.

MTI: writing review and editing.

MRI: Data analysis, writing review and editing.

MMR: GPS mapping, data analysis, writing review and editing.

OKI: Sortware, formal analysis, writing review and editing, resources.

IKJ: formal analysis, validation, writing review and editing, resources, supervision, project administration.

MAH: writing review and editing, supervision, project administration.

**Table S1:** Luna® Universal One-Step RT-qPCR Protocol

|  |  |  |
| --- | --- | --- |
| **PCR mix** |  |  |
| Reaction mix | : 10ul |  |
| RT-enzyme mix | : 1ul |  |
| S_F | : 0.8ul |  |
| S_R | : 1.2ul |  |
| Template | : 7ul |  |
| **PCR conditions** |  |  |
|  | Temperature | Time |
| RT | 55° C | 12 min |
| Initial denaturation | 95° C | 1 min |
| **Cycling part (40 cycles):** |  |  |
| Denaturation | 95° C | 30 sec |
| Annealing | 58° C | 45 sec |
| Extension | 60° C | 1 min |
| Final extension | 60° C | 7.5 min |
| Hold | 4° C | ∞ |
| **Gel Run** |  |  |
| Gel concentration | : 1% agarose |  |
| Gel volume | : 60ul |  |
| Dye | : Ethidium Bromide |  |
| Ladder | : 50bp , 6-7ul /well |  |
| PCR product | : 5ul/well |  |
| Voltage | : 80 V |  |
| Current | : 150 mA |  |
| Time | : 40 min |  |

**Table S2:** Sociodemographic profile and COVID 19 related information

| Variables | Percentage (%) | Frequency (n) |
| --- | --- | --- |
| **Sociodemographic profile** |  |  |
| Age |  |  |
| Less than 35 years | 9.3 | 10 |
| 35 to 60 years | 67.6 | 73 |
| More than 60 years | 23.2 | 25 |
| Sex |  |  |
| Male | 75.0 | 81 |
| Female | 25.0 | 27 |
| BMI |  |  |
| Normal | 49.1 | 53 |
| Overweight | 40.7 | 44 |
| Obese | 10.2 | 11 |
| Education |  |  |
| No education or primary incomplete | 14.0 | 15 |
| Secondary incomplete | 19.6 | 21 |
| Secondary or higher | 66.4 | 71 |
| Occupation |  |  |
| Unemployed/retired | 13.0 | 14 |
| Service/business | 60.2 | 65 |
| Homemaker | 18.5 | 20 |
| Others | 8.3 | 9 |
| Comorbidities |  |  |
| Yes | 50.0 | 54 |
| No | 50.0 | 54 |
| If yes, then are the comorbidities (Multiple response) |  |  |
| Asthma | 4.6 | 5 |
| Diabetes | 25.9 | 28 |
| High blood pressure | 31.5 | 34 |
| Low blood pressure | 3.7 | 4 |
| Cardiac disease | 9.3 | 10 |
| **Covid 19 related information** |  |  |
| Sanger variants |  |  |
| Unknown | 41.7 | 45 |
| UK | 5.6 | 6 |
| South Africa | 45.4 | 49 |
| Conventional and others | 7.41 | 8 |
| Symptoms (Multiple response) |  |  |
| Fever | 60.8 | 65 |
| Dry cough | 43.5 | 47 |
| Running nose | 11.1 | 12 |
| Muscle pain | 22.2 | 24 |
| Difficulties in breathing | 19.4 | 21 |
| Loss of smell | 25.9 | 28 |
| Loss of taste | 24.1 | 26 |
| Loss of appetite | 16.7 | 18 |
| Headache | 7.4 | 8 |
| Pain or swelling of legs, hands | 10.2 | 11 |
| Diarrhoea | 8.3 | 9 |
| Sore throat | 6.5 | 7 |
| Reason for covid 19 testing |  |  |
| Sick/hospitalized | 79.6 | 86 |
| Close contact with covid 19 patient | 8.3 | 9 |
| For travel and others | 12.0 | 13 |
| Outcome |  |  |
| Completely recovered | 63.0 | 68 |
| Still weak | 30.6 | 33 |
| Death | 6.5 | 7 |
| Complications after covid 19 positive (Multiple response) |  |  |
| Generalized weakness | 15.7 | 17 |
| Emotional changes | 7.4 | 8 |
| Dementia | 7.4 | 8 |
| Vision or changes in eye power | 0.9 | 1 |
| Loss of smell | 2.8 | 3 |
| Loss of taste | 3.7 | 4 |
| Muscle pain | 4.6 | 5 |
| Breathing problem | 1.9 | 2 |
| Vaccination |  |  |
| Yes | 38.9 | 42 |
| No | 61.1 | 66 |
| **Information on hospitalization** |  |  |
| Hospitalized |  |  |
| Yes | 15.7 | 17 |
| No | 84.3 | 91 |
| Duration |  |  |
| No hospitalization | 84.3 | 91 |
| 1 week | 9.3 | 10 |
| More than 1 week | 6.5 | 7 |
| ICU |  |  |
| Yes | 6.5 | 7 |
| No | 93.5 | 101 |
| Oxygen |  |  |
| Yes | 13.0 | 14 |
| No | 87.0 | 94 |

**Table S3:** Hospitalization in comorbid patients (including death cases)

|  | Immunized<br>% (n) | Non-immunized<br>% (n) | p-value |
| --- | --- | --- | --- |
| Duration |  |  |  |
| Discharged immediately | 76.5 (13) | 75.7 (28) | 1.000 |
| Hospitalized | 23.5 (4) | 24.1 (9) |  |

\*No significant difference was observed. Sample size is too small to draw any decision.

**Table S4:** Whole genome sequence analysis among immunized and unimmunized group for the confirmation of SARS-CoV-2 variants.

| WGS ID | Accession no | Vaccination history | Disease outcome | Variant | synonymous mutation | Non-synonymous mutation |
| --- | --- | --- | --- | --- | --- | --- |
| w15 | EPI_ISL_2350167 | Completely immunized | Recovered | 20B | 14 | 7 |
| w4 | EPI_ISL_2348653 | Partially Immunized | Death | SA (Beta V2) | 36 | 11 |
| w7 | EPI_ISL_2348658 | Partially Immunized | Recovered | SA (Beta V2) | 32 | 22 |
| w8 | EPI_ISL_2348660 | Partially Immunized | still weak | SA (Beta V2) | 33 | 19 |
| w9 | EPI_ISL_2348661 | Partially Immunized | still weak | SA (Beta V2) | 28 | 21 |
| w10 | EPI_ISL_2348662 | Partially Immunized | Recovered | NY (Eta) | 35 | 13 |
| w3 | EPI_ISL_2349879 | Partially Immunized | Recovered | 20B | 20 | 13 |
| w12 | EPI_ISL_2348664 | Unimmunized | Recovered | SA (Beta V2) | 20 | 22 |
| w13 | EPI_ISL_2349692 | Unimmunized | Death | SA (Beta V2) | 33 | 11 |
| w6 | EPI_ISL_2350142 | Unimmunized | Recovered | SA (Beta V2) | 30 | 20 |
| w2 | EPI_ISL_2348652 | Unimmunized | Recovered | B.1.1.7 (Alpha V1) | 24 | 34 |
| w11 | EPI_ISL_2348663 | Unimmunized | Recovered | NY (Alpha V1) | 30 | 22 |
| w5 | EPI_ISL_2349880 | Unimmunized | Death | Conventional (Alpha V1) | 32 | 22 |
| w1 | EPI_ISL_2348651 | Unimmunized | still weak | 20B | 22 | 12 |

**Figure S1:**
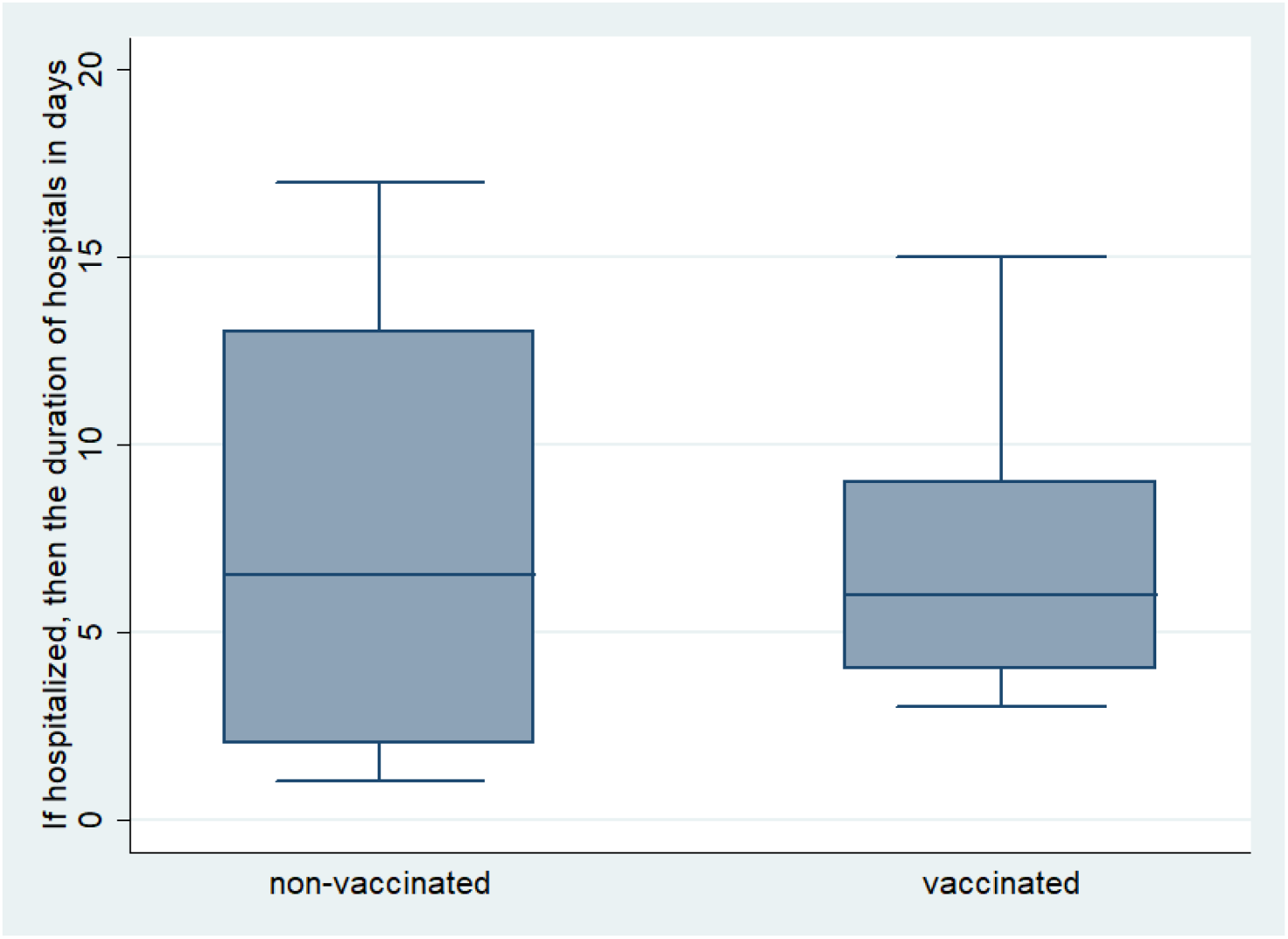
Comparison of the length of hospitalization among the immunized (12%, n=5) and unimmunized (18%, n=12) severe SARS-COV-2 infected patients (p=0.383).

**Figure S2:**
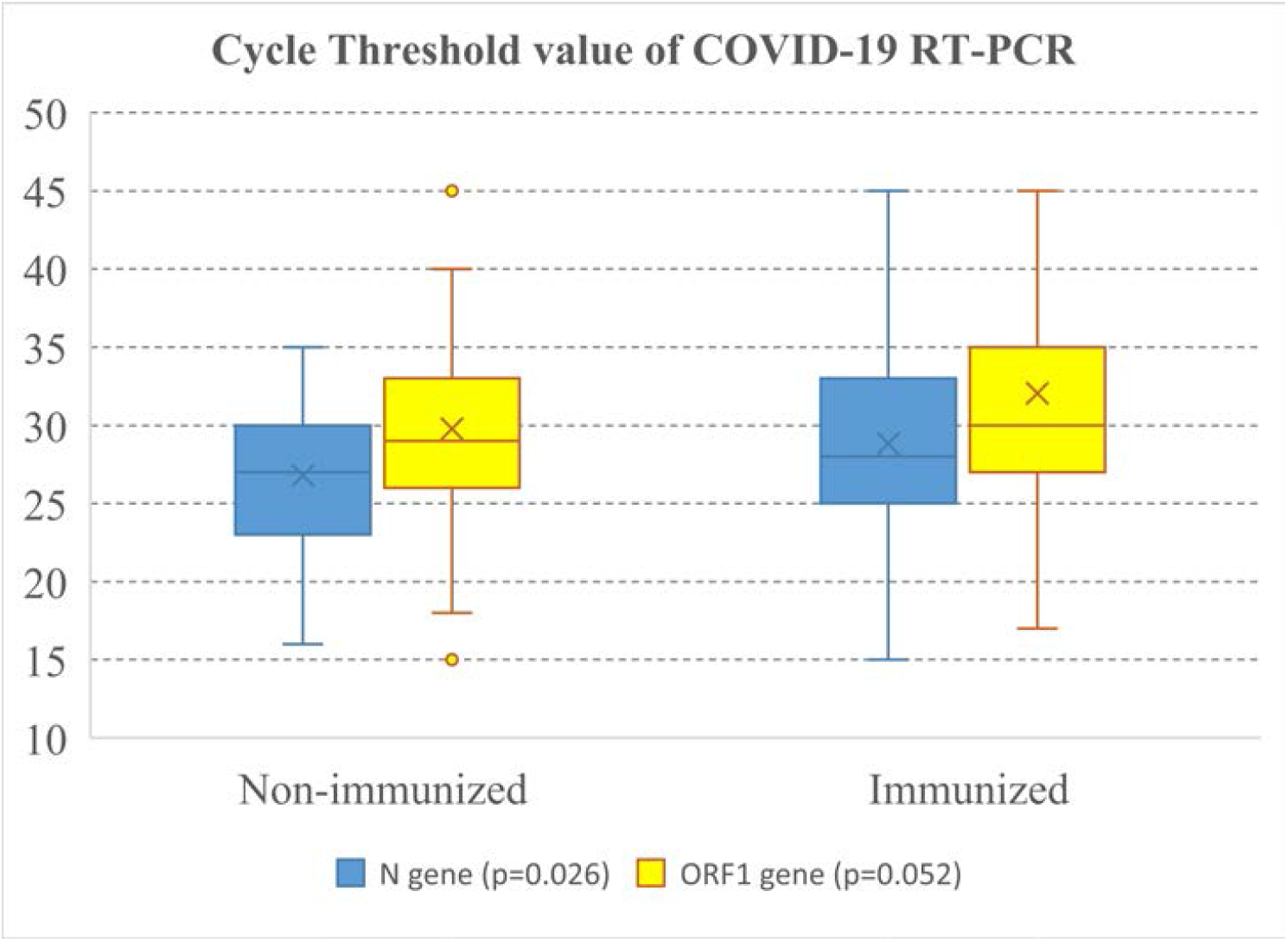
The differences of *N*-gene and *ORF1b*-gene cycle threshold values among immunized and unimmunized group during the detection of COVID-19 using RT-PCR kit. The unamplified gene products were considered with a CT value 45, the maximum cycle of the experiment. The independent mean t-test were performed for N-gene (p=0.026) and ORF1 gene (p=0.052).

